# Delayed cerebrovascular reactivity in individuals with spinal cord injury in the right inferior parietal lobe: a breath-hold functional near-infrared spectroscopy study

**DOI:** 10.1101/2024.06.03.24307819

**Authors:** Donna Y. Chen, Xin Di, Keerthana Deepti Karunakaran, Hai Sun, Saikat Pal, Bharat B. Biswal

**Affiliations:** Department of Biomedical Engineering, New Jersey Institute of Technology, Newark, NJ, US; Rutgers Biomedical and Health Sciences, Rutgers School of Graduate Studies, Newark, NJ, US; Boston Children’s Hospital, Harvard Medical School, Boston, MA, US; Department of Neurosurgery, Rutgers Robert Wood Johnson Medical School, New Brunswick, NJ, US; Electrical and Computer Engineering Department, New Jersey Institute of Technology, Newark, NJ, US; Spinal Cord Damage Research Center, James J. Peters Veterans Affairs Medical Center, Bronx, NY, US

## Abstract

Cerebrovascular reactivity (CVR) reflects the ability of blood vessels to dilate or constrict in response to a vasoactive stimulus, and allows researchers to assess the brain’s vascular health. Individuals with spinal cord injury (SCI) are at an increased risk for autonomic dysfunction in addition to cognitive impairments, which have been linked to a decline in CVR; however, there is currently a lack of brain-imaging studies that investigate how CVR is altered after SCI. In this study, we used a breath-holding hypercapnic stimulus and functional near-infrared spectroscopy (fNIRS) to investigate CVR alterations in individuals with SCI (n = 20, 14M, 6F, mean age = 46.3 ± 10.2 years) as compared to age– and sex-matched able-bodied (AB) controls (n = 25, 19M, 6F, mean age = 43.2 ± 12.28 years). CVR was evaluated by its amplitude and delay components separately by using principal component analysis and cross-correlation analysis, respectively. We observed significantly delayed CVR in the right inferior parietal lobe in individuals with SCI compared to AB controls (linear mixed-effects model, fixed-effects estimate = 6.565, Satterthwaite’s t-test, t = 2.663, p = 0.008), while the amplitude of CVR was not significantly different. The average CVR delay in the SCI group in the right inferior parietal lobe was 14.21 s (sd: 6.60 s), and for the AB group, the average delay in the right inferior parietal lobe was 7.08 s (sd: 7.39 s). CVR delays were also associated with the duration since injury in individuals with SCI, in which a longer duration since injury was associated with a shortened delay in CVR in the right inferior parietal region (Pearson’s r-correlation, r = –0.59, p = 0.04). This study shows that fNIRS can be used to quantify changes in CVR in individuals with SCI, and may be further used in rehabilitative settings to monitor the cerebrovascular health of individuals with SCI.

## 1. Introduction

Spinal cord injury (SCI) is caused by damage to the spinal cord that can be either traumatic or non-traumatic, and affects over 15 million individuals world-wide (World Health Organization, 2024). Traumatic cases of SCI are caused by external physical forces that damage the spinal cord whereas non-traumatic cases of SCI are typically caused by chronic disease processes such as degenerative disc disease (Ahuja et al., 2017). In both cases, SCI leads to socioeconomic burdens on the individuals and increased difficulties with daily tasks, causing psychological distress in many individuals (Craig et al., 2009). Following a primary spinal cord injury event, secondary injury mechanisms occur, such as the increase in neuroinflammation (David & Kroner, 2011; Donnelly & Popovich, 2008; Faulkner et al., 2004; Hellenbrand et al., 2021), increased oxidative stress (Anwar et al., 2016; Carlson et al., 1998), vascular disruption (Siddiqui et al., 2015; Sinescu et al., 2010), cell death (Couillard-Despres et al., 2017), and autonomic dysfunction (Alizadeh et al., 2019). These secondary injury mechanisms often cause further neurological damage and health complications (Silva et al., 2014). Therefore, it is critical to investigate chronic injury mechanisms and monitor potential secondary health conditions to improve the quality of life for individuals with SCI.

A particular secondary health concern for individuals with SCI is the risk of autonomic dysfunction, which could lead to ischemic stroke and autonomic dysreflexia (Banerjea et al., 2008; Cragg et al., 2013; LaVela et al., 2012; Wu et al., 2012). After injury to the spinal cord, afferent and efferent nerve fibers of the autonomic nervous system may be severed, resulting in altered autonomic nervous system function due to disrupted supraspinal pathways (Henke et al., 2022). This in turn causes impairments in cardiovascular function, which contributes to reduced function in brain perfusion, as well as cerebral autoregulation, cerebrovascular reactivity, and neurovascular coupling (Kim & Tan, 2018). In particular, cerebrovascular reactivity (CVR) has been widely used as a tool to investigate the state of vascular health in the brain (Sleight et al., 2021). CVR reflects how well blood vessels can constrict or dilate in response to a vasoactive stimulus, and can be measured using different tools, such as using functional magnetic resonance imaging (fMRI) coupled with hypercapnic or hypocapnia stimuli (Bright & Murphy, 2013; Di et al., 2013; Fierstra et al., 2013; Kastrup et al., 1999, 2001; Pinto et al., 2021). Reduced CVR has been associated with cognitive impairments, which has been commonly reported after SCI, with individuals with SCI having a 13 times greater risk for cognitive impairments than able-bodied individuals (Alcántar-Garibay et al., 2022; Craig et al., 2017; Davenport et al., 2012; Nightingale et al., 2020; Sachdeva et al., 2018). However, there is currently a lack of studies investigating how CVR is altered after SCI.

Weber and colleagues used fMRI to investigate CVR in individuals with SCI via administration of CO2 gas and found increased CVR response time (delay) in individuals with SCI compared to that of able-bodied controls (Weber et al., 2022). However, they found no significant differences in the CVR metric on its own when it was not separated into its steady-state and active components. They also report significant associations between the time-since-injury and the neurological level of injury of the SCI participants and the steady-state CVR component; however, the authors note the limitations in sample size of 7 SCI and 6 controls in their study (Weber et al., 2022). This shows that CVR changes with different injury characteristics and specific components of CVR are altered after one sustains an injury to the spinal cord. It is important to note that there are practical issues with getting individuals with SCI into an MRI scanner, such as the possibility of having metallic implants that are not MRI-compatible, difficulties with placing individuals from the wheelchair to the supine position in the MRI scanner, and potential discomfort in the rigid supine position in the MRI scanner, particularly for those who may feel claustrophobic (Blight et al., 2019; Weber et al., 2022).

The use of functional near-infrared spectroscopy (fNIRS) may resolve some of these issues in measuring brain hemodynamic activity in individuals with SCI. fNIRS is a relatively comfortable device placed on the scalp that uses near-infrared light to measure brain hemodynamic activity, similar to that of fMRI, based on the optical properties of deoxygenated and oxygenated hemoglobin in the cortical brain area (Jöbsis, 1977; Delpy et al., 1988; Villringer et al., 1993; Yücel et al., 2017). Despite its lower spatial resolution, it has a higher temporal resolution than fMRI (Ferrari & Quaresima, 2012), up to 100 Hz, which offers an advantage in better characterizing the delays in CVR (Amyot et al., 2020; MacIntosh et al., 2003). fNIRS has been previously used to quantify CVR in both breath-holding and gas inhalation settings (Amyot et al., 2020, 2022; Karunakaran et al., 2021; S. Miller & Mitra, 2021; Reddy et al., 2021; Smielewski et al., 1995; Vagné et al., 2020). Wilson and colleagues measured CVR in 6 individuals with tetraplegia using both transcranial doppler and fNIRS, and reported similar CVR in response to CO2 in tetraplegia and able-bodied control groups, suggesting cerebrovascular adaptation with chronic tetraplegia (Wilson et al., 2010). It is not clear whether CVR as measured by fNIRS is altered in individuals with chronic paraplegia, as the level of injury and completeness of injury can impact the degree of autonomic dysfunction (Henke et al., 2022; West et al., 2013).

In the present study, we investigated both the amplitude and delay of CVR in SCI and able-bodied (AB) groups, as measured by fNIRS during a hypercapnic breath-hold task. We used principal component analysis (PCA) and cross correlation analysis (CCA) to determine the amplitude and delays associated with the breath-hold CVR response, respectively. These CVR metrics were obtained for all subjects and then compared between the AB and SCI groups, across cognitive and motor brain regions, for all hemoglobin species. We then investigated the association between CVR characteristics and injury characteristics in the SCI group for particular brain regions which revealed differences between the AB and SCI groups. By studying CVR in individuals with SCI, we can better understand secondary injury mechanisms which occur after spinal cord injury and develop more effective rehabilitative treatments.

## 2. Materials and Methods

### 2.1. Participants

All participants were recruited from the New Jersey/New York metropolitan area. A total of 20 participants with SCI (14M, 6F, mean age = 46.3 ± 10.2 years) and 25 age– and sex-matched able-bodied (AB) controls (19M, 6F, mean age = 43.2 ± 12.28 years) participated in this study. Institutional Review Board (IRB) approval was obtained, and all participants provided written informed consent to the study. Participants were excluded from the study if any of the following applied to them: (1) within one-year post-spinal cord injury or in the acute phase of injury, (2) presence of tetraplegia or inability to perform upper limb motor movements, (3) history of or concurrent traumatic brain injury (TBI), (4) history of or concurrently has psychiatric disorders such as post-traumatic stress disorder, addiction, bipolar disorder, or schizophrenia, (5) presence of acute illness or infection, (6) history of chronic hypertension, diabetes mellitus, stroke, epilepsy or seizure disorders, multiple sclerosis, Parkinson’s disease, (7) illicit drug abuse within the past 6 months, (8) has Alzheimer’s disease or dementia, (9) not able to speak English, and (10) younger than 18 years of age or older than 65 years of age (D. Y. Chen, Di, Amaya, et al., 2024). The level of spinal injury, completeness of injury, and the duration since injury were collected from participants with SCI (Table 1).

**Table 1.** Demographic information and clinical characteristics from participants with SCI enrolled in the current study.

| ID | Age (years) | Sex | Spinal Level of Injury | # Spinal Segments Affected | Complete (C) /Incomplete (I) | Duration since Injury (years) | Handedness |
| --- | --- | --- | --- | --- | --- | --- | --- |
| SCI-01 | 45-61 | M | T11 | 11 | C | 12 | Right |
| SCI-02 | 45-61 | M | T2 | 20 | C | 16 | Left |
| SCI-03 | 45-61 | M | T10/11 | 12 | C | 6 | Right |
| SCI-04 | 28-44 | M | T12 | 10 | C | 9 | Right |
| SCI-05 | 28-44 | M | T11 | 11 | C | 14 | Right |
| SCI-06 | 45-61 | M | T4 | 18 | C | 10 | Right |
| SCI-07 | 28-44 | F | T4 | 18 | I | 8 | Right |
| SCI-08 | 28-44 | F | L1 | 9 | I | 12 | Left |
| SCI-09 | 45-61 | M | T6 | 16 | C | 24 | Right |
| SCI-10 | 28-44 | F | T11 | 11 | C | 26 | Left |
| SCI-11 | 45-61 | M | T11/T12 | 11 | C | 26 | Right |
| SCI-12 | 45-61 | M | T1 | 21 | I | 27 | Right |
| SCI-13 | 45-61 | M | C5 | 24 | I | 30 | Right |
| SCI-14 | 28-44 | M | T6 | 16 | I | 2 | Right |
| SCI-15 | 45-61 | F | L1 | 9 | I | 30 | Right |
| SCI-16 | 45-61 | M | T7 | 15 | I | 30 | Right |
| SCI-17 | 45-61 | M | T7 | 15 | C | 10 | Right |
| SCI-18 | 45-61 | F | C4/C5 | 25 | I | 10 | Left |
| SCI-19 | 45-61 | M | T4 | 18 | C | 16 | Right |
| SCI-20 | 28-44 | F | C5/C6 | 24 | I | 11 | Left |

### 2.2. Breath-hold Task Paradigm

All participants performed a breath-hold task paradigm consisting of 6 blocks of alternating periods of 15 s breath-hold and 30 s resting periods (Figure 1A). The task started off with 30 s of rest initially, during which the participants were instructed to breathe at their own comfortable pace in a relaxed manner. All instructions were displayed via a computer screen using E-Prime 2.0 software (Psychology Software Tools, Pittsburgh, PA), instructing the participants when to hold their breath and when to breathe normally. The words “rest” and “hold” were displayed in white font at the center of the screen on a black background. The participants also performed a resting-state task and N-back working memory task as part of a larger study; however, due to the focus of this study being cerebrovascular reactivity, only the breath-hold task results are analyzed and discussed here.

**Figure 1.**
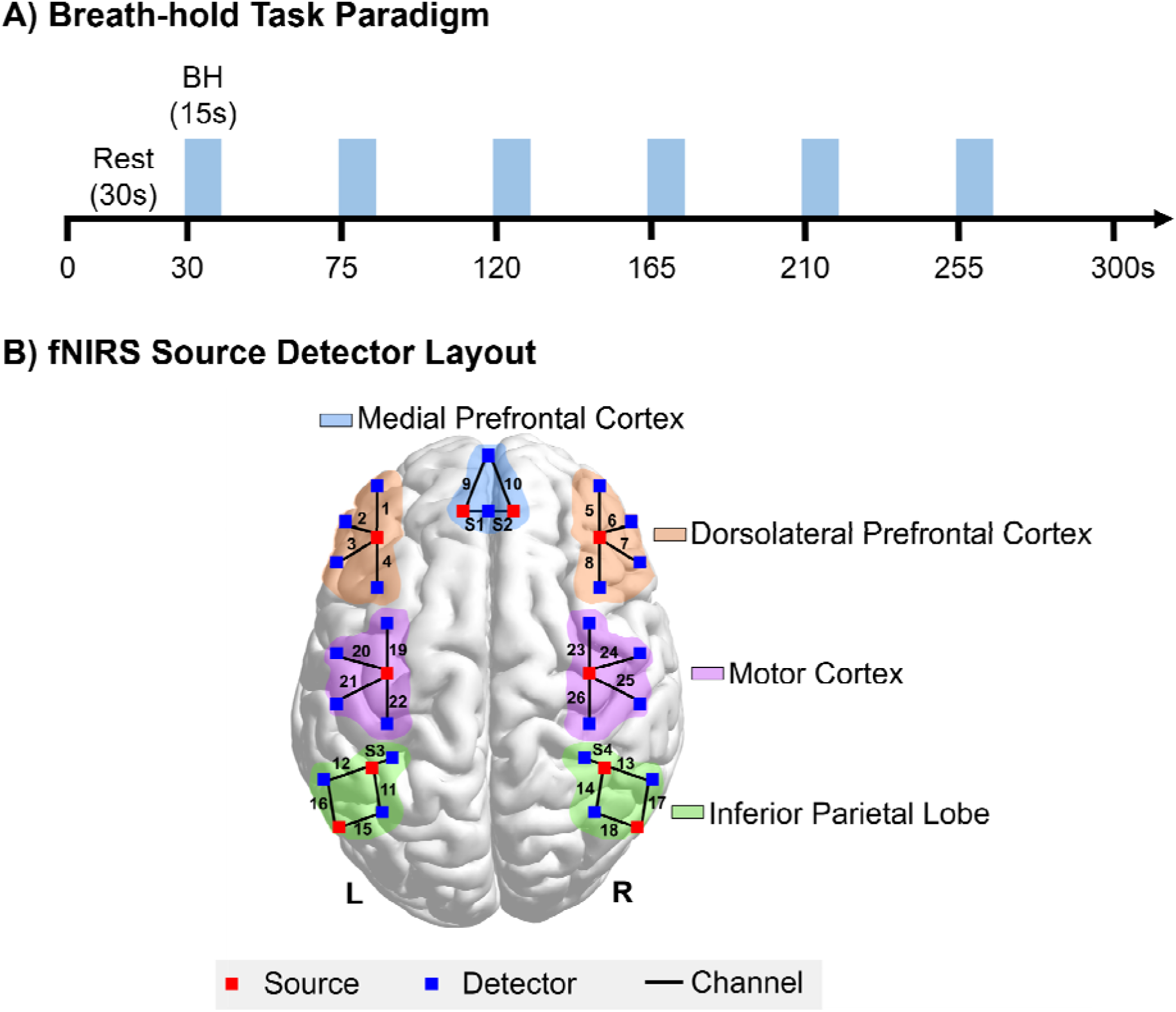
**A**) fNIRS Breath-hold (BH) task paradigm consisting of 6 breath-hold sessions, 15s each, interleaved with 30s of rest. **B)** fNIRS Optode layout with 10 optode sources and 24 detectors configured across the medial prefrontal cortex, dorsolateral prefrontal cortex, motor cortex, and inferior parietal lobe. A total of 26 channels are used with a source-detector distance of 30mm and 4 short separation channels are used with a source-detector distance of 8.4mm, as denoted by S1, S2, S3, and S4.

### 2.3. fNIRS Data Acquisition

A continuous wave fNIRS system was used for all fNIRS data acquisition at a sampling frequency of 25 Hz, using both 690 nm and 830 nm of light (CW6 System, TechEn Inc., Milford, MA). A total of 10 optode sources and 24 detectors were arranged across the head to construct 26 long source-detector separation channels and 4 short channels (Figure 1B). The long channels were placed at a source to detector distance of 30 mm while the short channels were placed at a source to detector distance of 8.4 mm (Brigadoi & Cooper, 2015). These short channels were placed bilaterally on the left and right inferior parietal lobes and in the medial prefrontal cortex to reduce the impact of physiological noise from extracerebral regions on the data of interest. The 26 long channels covered the following regions of interest (ROI): the left and right dorsolateral prefrontal cortex (LDLPFC, RDLPFC, channels 1-4 and channels 5-8, respectively), medial prefrontal cortex (MPFC, channels 9&10), left and right motor cortex (LMOTOR, RMOTOR, channels 19-22 and channels 23-26, respectively), and left and right inferior parietal lobes (LIPL, RIPL, channels 11, 12, 15, & 16, and channels 13, 14, 17, & 18, respectively). The ROI were identified on each participant’s head using a neural navigation system (Brain Sight Neural Navigator, Rogue Research Inc., Canada) with the Montreal Neurological Institute’s (MNI-152) average structural brain template as a reference.

### 2.4. fNIRS Data Preprocessing

The fNIRS data were preprocessed in the following order: (1) raw light intensity values were converted to optical density values, (2) wavelet-based head-motion correction was performed using a threshold of 1.5 times the interquartile range (Molavi & Dumont, 2012), (3) band-pass filtering was applied using a pass-band from 0.01 to 0.15 Hz, (4) data were then converted to oxygenated hemoglobin (HbO), deoxygenated hemoglobin (HbR), and total hemoglobin (HbT) using the modified Beer-Lambert law (Delpy et al., 1988), (5) short channel regression was applied, and (6) channels were excluded based on the relative power in the task frequency range (Figure 2). Short channel regression was implemented using the proximal channel method, in which a short channel regressor was chosen for each long channel based on its proximity to the long channel (Gagnon et al., 2012). In this case, channel S1 was used to regress out noise from channels 1-4, & 9, channel S2 was used to regress out noise from channels 5-8, & 11, channel S3 was used to regress out noise from channels 11, 12, 15, 16, & 19-22, and channel S4 was used to regress out noise from channels 13, 14, 17, 18, & 23-26 (Figure 2). The short channel regression models were solved using the ordinary least squares method and were formatted as

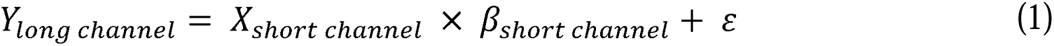

**Figure 2.**
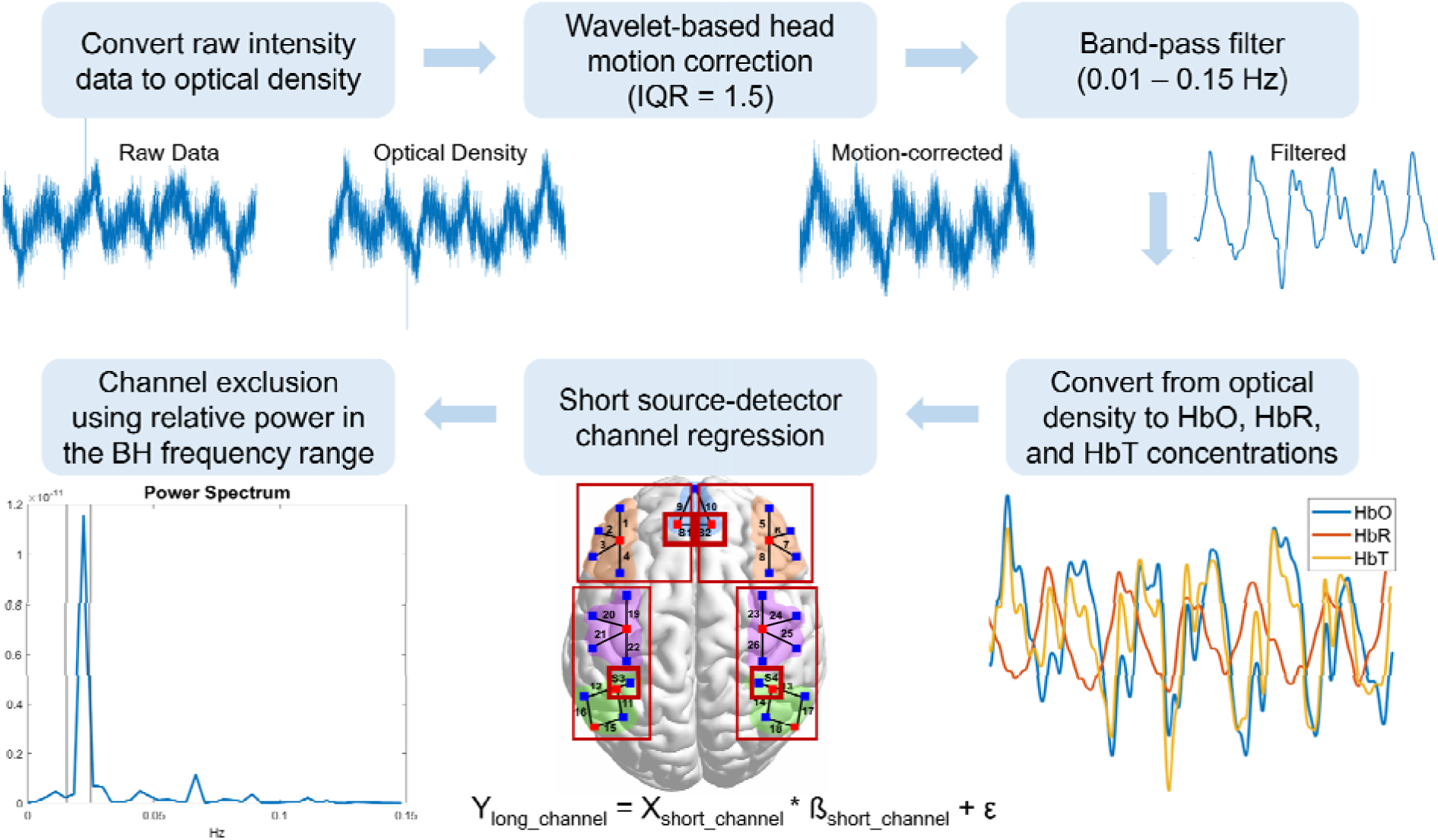
fNIRS preprocessing pipeline applied to both SCI and AB groups. The preprocessing steps were carried out in the following order: 1) convert raw intensity data to optical density data, 2) perform wavelet-based head motion correction with 1.5 times the interquartile range threshold, 3) band-pass filter from 0.01 to 0.15 Hz, 4) convert the filtered optical density data to oxy-hemoglobin (HbO), deoxy-hemoglobin (HbR), and total-hemoglobin (HbT) data, 5) regress the short source-detector channel from the long channels, and 6) perform channel exclusion using the relative power in the breath-hold frequency range. The short source-detector channel regression and channel exclusion steps were applied to all hemoglobin species.

where Y_long channel_ represents the fNIRS data from one of the long channels, X_short channel_ represents the fNIRS data from the proximal short channel, and ε represents the residuals. Short channel regression was performed on all subject data, across each channel separately and the residuals were kept for further analyses (Eq. 1).

After short channel regression, fNIRS channels were excluded based on the relative power in the breath-hold frequency range. Since the breath-hold task period occurred every 45 s (15 s breath-hold and 30 s rest), the dominant breath-hold task frequency range was defined as 40 s to 65 s, or 0.0154 Hz to 0.025 Hz. Therefore, the relative power was computed as the average power in the breath-hold frequency range (0.0154 to 0.025 Hz) divided by the total average power in the frequency range from 0 to 12 Hz (up to the Nyquist frequency) (Eq. 2).

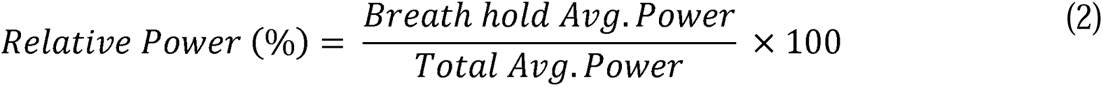

The average power was computed using the *bandpower* function in MATLAB 2024a (The MathWorks Inc., Natick, Massachusetts). Channels with less than 50% relative power in the breath-hold frequency range were considered “insufficient” and were excluded from subsequent analyses (Zvolanek et al., 2023). This method of channel exclusion not only allows us to remove noisy data, but also offers a measure of task compliance, since those who did not comply with the task would likely not show high power in the breath-hold frequency range. Short channel regression and channel exclusion were applied to HbO, HbR, and HbT separately, and sufficient data from each hemoglobin species were analyzed in the subsequent steps. Data from the start of the breath-hold task (beginning of the first 15s breath-hold period) to the end of the entire task (including the last 30s rest period) were analyzed. The first 30s of rest were discarded since it was included for the participants to relax and serve as a buffer period before the breath-hold stimuli began. All data preprocessing scripts were written and executed in MATLAB 2024a (The MathWorks Inc., Natick, Massachusetts) and used HOMER3 functions (Huppert et al., 2009).

### 2.5. Principal Component Analysis

After the fNIRS data were preprocessed, all data were standardized to z-scores by subtracting each timepoint by the mean of the entire time-series and then dividing by the standard deviation. Z-score standardization was performed for all channels and all subjects, for each hemoglobin species concentration separately. Principal component analysis (PCA) using a singular value decomposition algorithm was then applied to the z-scored data for each channel, in which the data from subjects in both AB and SCI groups were aggregated together in a [timepoint x subject] matrix. The square root of the eigenvalues of the covariance matrix was taken and multiplied by the eigenvector to obtain the principal component loadings. Since the first principal component explained the most variance, the coefficients of the first principal component (PC1 loading) were analyzed further. The PC1 loading value was inferred to represent the breath-hold response and serves as a measure of CVR amplitude, in which a higher PC1 loading represents a greater CVR amplitude response. PCA is a model-free method that allows us to forgo *a priori* assumptions on the shape and delay of the hemodynamic response function (HRF) (Di & Biswal, 2022; Hejnar et al., 2007), thus offering a more unbiased measure of the breath-hold CVR amplitude response. The PC1 loading values were compared between the AB and SCI groups for each brain region and each hemoglobin species.

### 2.6. Breath-hold CVR Response Delays

In addition to obtaining a measure of CVR amplitude, we investigated differences in the delays of the breath-hold CVR response by performing cross-correlation analysis. Cross-correlation analysis was applied between the preprocessed fNIRS data and the task design convolved with the canonical HRF model (Friston et al., 1998). The fNIRS time-series data were shifted by a delay range of 0 s to 20 s in increments of 0.04 s (sampling frequency of 25 Hz) with respect to the convolved task design data. This delay range was chosen based on the physiological range of breath-hold delays that has been reported in the literature (K. Chen et al., 2021; Gong et al., 2023; Moia et al., 2020; Stickland et al., 2022; Tong et al., 2011). The “optimal” delay was chosen based on the delay or lag value that yielded the highest correlation value when shifting the time-series data. The optimal delay was calculated for each subject, each channel, and each hemoglobin species. These delay values were then compared between the AB and SCI groups across all brain regions.

### 2.7. CVR and Injury Characteristics

After comparing the CVR amplitude and delay values between the AB and SCI groups, we further investigated whether injury characteristics in the SCI group were associated with CVR characteristics. To reduce the number of comparisons, injury characteristics were correlated with PC1 loading and delay values for any regions that showed significant differences between the AB and SCI group. For the SCI group, we investigated the duration since injury (in years) and the level of spinal injury score, calculated as the number of spinal segments affected that are below the level of injury (Table 1). The mean of the CVR amplitude and delay values were taken across all channels within the particular ROI to obtain a single ROI-specific CVR metric for each subject. The mean value was taken since not all subjects had data from all channels within a particular ROI, due to the channel pruning procedure implemented.

### 2.8. Statistical Analysis

We compared CVR amplitude and delays between the SCI and AB groups using mixed-effects modeling. The linear mixed-effects models were implemented in R using the *lme4* package (Bates et al., 2015) and were formatted as follows:

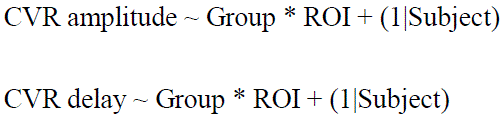

Linear mixed effects models were applied to all HbO, HbR, and HbT concentration data separately. The “Group” variable consisted of either the SCI or AB category and the “ROI” variable consisted of the 7 ROI across the fNIRS cap: the LDLPFC, RDLPFC, MPFC, LMOTOR, RMOTOR, LIPL, and RIPL regions. The “Group” and “ROI” variables were treated as fixed effects while the “Subject” variable was treated as a random effect. Not all subjects had data for all channels within a particular ROI due to the channel pruning procedure. Therefore, by implementing a mixed-effects modeling approach, we were able to account for the differences in the number of channels with sufficient data quality from each subject. All linear mixed effects models were fit by using the restricted maximum likelihood algorithm and t-tests were performed using Satterthwaite’s method (Kuznetsova et al., 2017). For analysis of associations between CVR amplitude and delays with the injury characteristics in the SCI group, Pearson’s r correlation was used for each hemoglobin species concentration.

## 3. Results

For our quality control analyses, after preprocessing all fNIRS data, the relative power in the breath-hold frequency range was calculated for all HbO, HbR, and HbT concentrations, across all subjects and channels. High relative power values were observed across Channels 1-10, while lower relative power values were observed in Channels 11-26, across all hemoglobin concentrations (Figure 3 A-C). The data included in subsequent analyses and determined as “sufficient” were consistent across hemoglobin concentrations and consistent between the AB and SCI groups (Figure 3 D-F).

**Figure 3.**
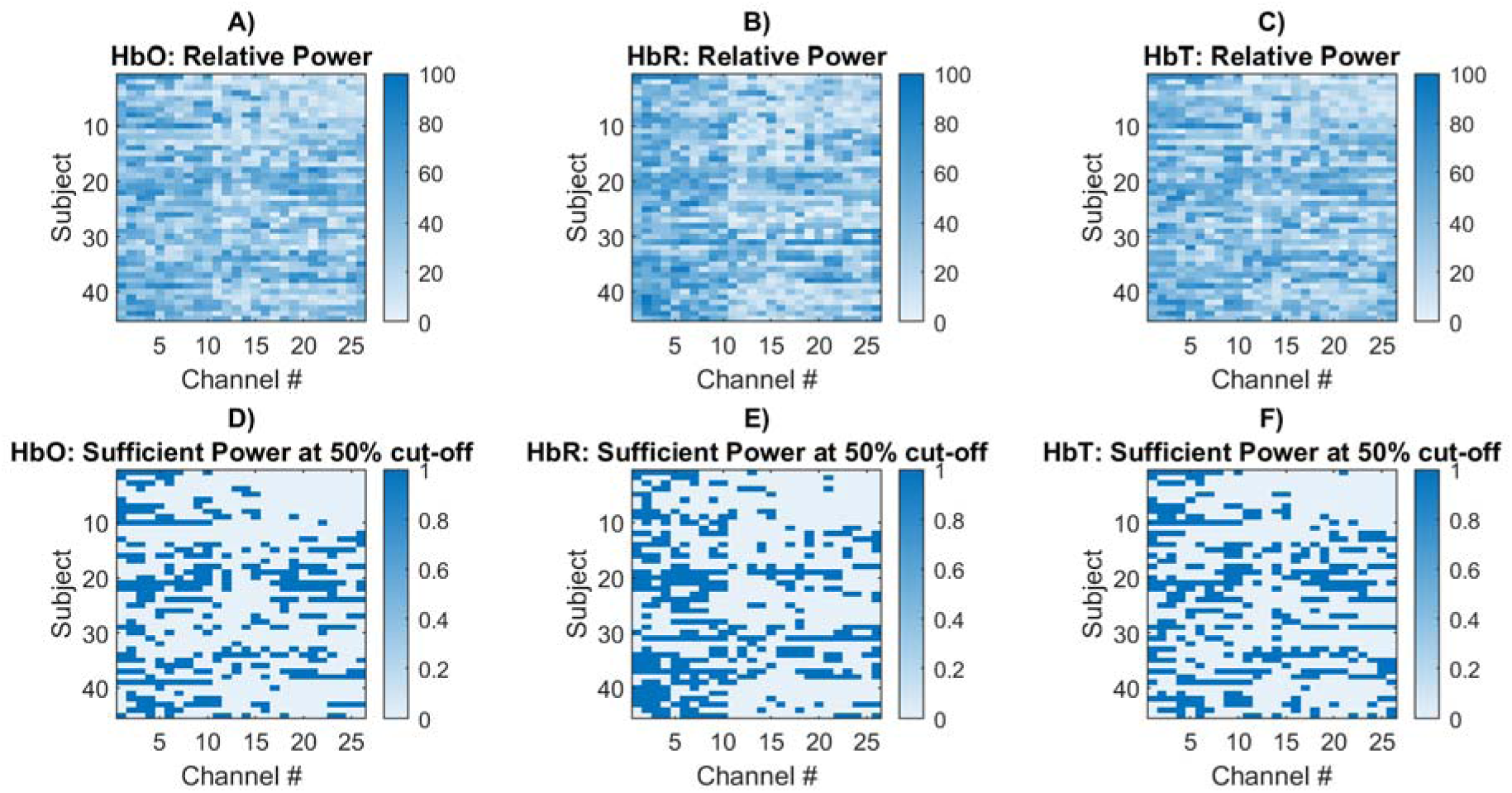
Relative power in the breath-hold frequency range shown for each subject across all channels, shown for. **A)** oxygenated hemoglobin (HbO), **B)** deoxygenated hemoglobin (HbR), and **C)** total hemoglobin (HbT). Subjects 1-25 are individuals in the AB group whereas subjects 26-45 are individuals in the SCI group. After applying a threshold of 50% relative power, channels that were included are indicated by 1’s and channels that were excluded are indicated by 0’s, for **D)** HbO, **E)** HbR, and **F)** HbT.

### 3.1. CVR Amplitude in SCI vs. AB

Across all brain regions, there were no significant differences in CVR amplitude, as measured by PC1 Loading scores, between the SCI and AB groups (Figure 4A-C). Separate linear mixed effects models were generated for each hemoglobin concentration species and no significant differences in CVR amplitude were observed between SCI and AB groups across HbO, HbR, and HbT (Table 2). For HbT, although the AB group showed higher CVR amplitude in the right motor region, the effect was weak (fixed-effect estimate = –0.187, t = –2.214, p = 0.028). Overall, the PC1 Loading values hovered around 0.1 for both SCI and AB groups and showed similar values across hemoglobin concentration species (Figure 4).

**Figure 4.**
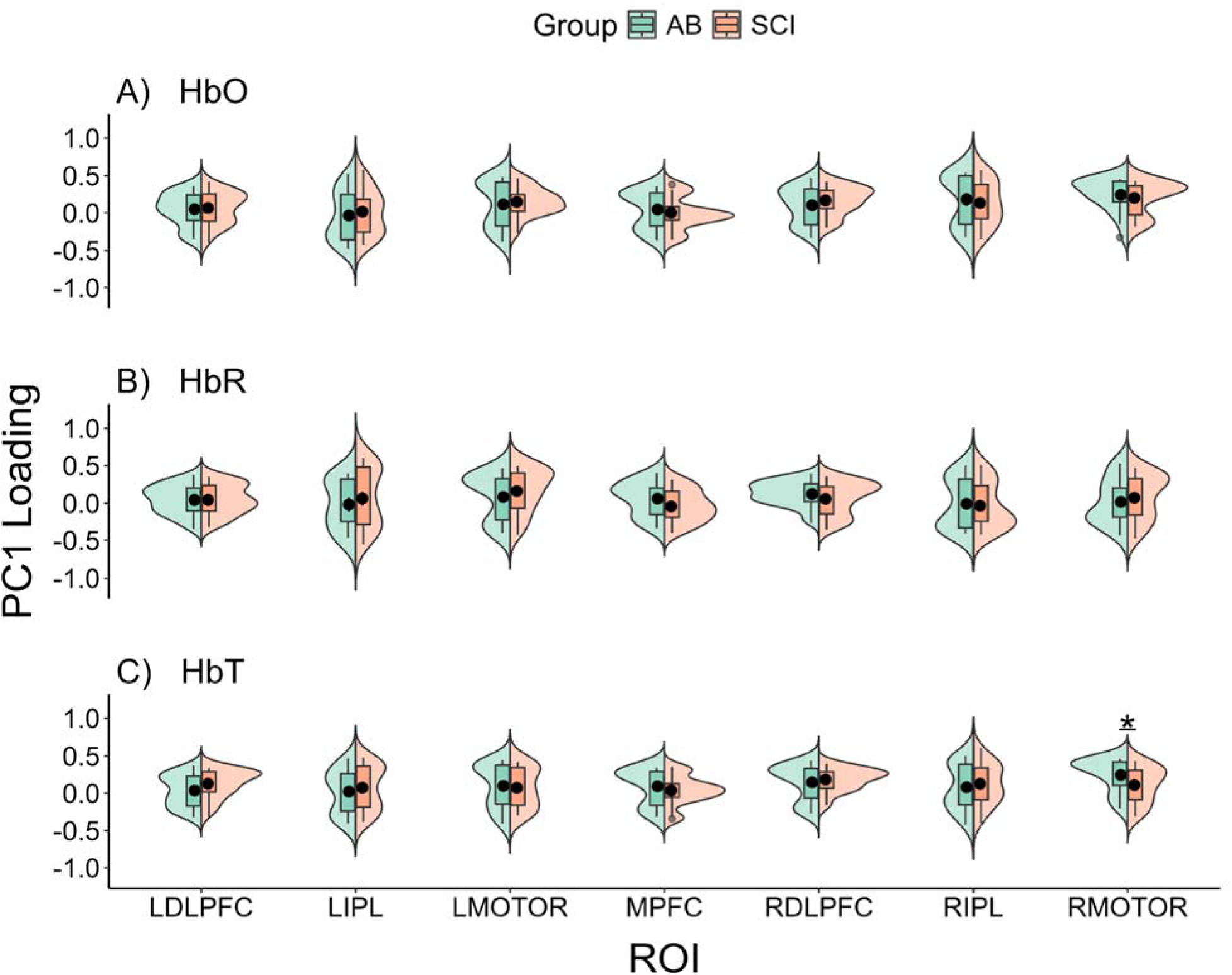
PC1 Loadings shown for both AB and SCI groups, for all hemoglobin species: **A)** oxy-hemoglobin (HbO), **B)** deoxy-hemoglobin (HbR), and **C)** total hemoglobin (HbT), across all brain regions of interest: left dorsolateral prefrontal cortex (LDLPFC), left inferior parietal lobe (LIPL), left motor cortex (LMOTOR), medial prefrontal cortex (MPFC), right dorsolateral prefrontal cortex (RDLPFC), right inferior parietal lobe (RIPL), and right motor cortex (RMOTOR). The means of the PC1 Loadings are indicated by black circles and the distribution of the AB and SCI groups are shown, denoted by green and orange, respectively.

**Table 2.** Mixed effects model results for CVR amplitude comparisons between SCI and AB groups, across each ROI. One ROI is left out in the reports (the LDLPFC region) due to being the reference level for all comparisons.

| ROI | Estimate |  |  | Standard Error |  |  | t-value |  |  | p-value |  |  |
| --- | --- | --- | --- | --- | --- | --- | --- | --- | --- | --- | --- | --- |
|  | HbO | HbR | HbT | HbO | HbR | HbT | HbO | HbR | HbT | HbO | HbR | HbT |
| LIPL | -0.007 | 0.086 | -0.044 | 0.096 | 0.108 | 0.083 | -0.070 | 0.792 | -0.531 | 0.944 | 0.429 | 0.596 |
| LMOTOR | 0.044 | 0.061 | -0.099 | 0.095 | 0.098 | 0.085 | 0.468 | 0.616 | -1.176 | 0.640 | 0.538 | 0.241 |
| MPFC | -0.081 | -0.113 | -0.102 | 0.102 | 0.096 | 0.090 | -0.793 | -1.187 | -1.135 | 0.429 | 0.236 | 0.257 |
| RDLPFC | 0.072 | -0.048 | 0.004 | 0.085 | 0.076 | 0.077 | 0.839 | -0.630 | 0.057 | 0.402 | 0.529 | 0.955 |
| RIPL | -0.029 | -0.034 | -0.014 | 0.105 | 0.096 | 0.086 | -0.275 | -0.350 | -0.164 | 0.783 | 0.726 | 0.870 |
| RMOTOR | -0.041 | 0.037 | -0.187 | 0.097 | 0.096 | 0.085 | -0.420 | 0.387 | -2.214 | 0.675 | 0.699 | 0.028<br>* |

### 3.2. CVR Delays in SCI vs. AB

We observed significantly longer CVR delays in the SCI group compared to the AB group in the right inferior parietal lobe, for HbR concentration (linear mixed-effects model, fixed-effects estimate = 6.565, Satterthwaite’s t-test, t = 2.663, p = 0.008) (Table 3). The average HbR CVR delay in the SCI group in the right inferior parietal lobe was 14.21 s (sd: 6.60 s), and for the AB group, the average delay in the right inferior parietal lobe was 7.08 s (sd: 7.39 s) (Figure 5). For HbO concentration, we observed slightly higher CVR delay values in the left inferior parietal lobe for the AB group compared to the SCI group (linear mixed-effects model, fixed-effects estimate = –6.027, Satterthwaite’s t-test, t = –2.518, p = 0.012) (Table 3), with the mean delay values in the AB group being 9.53 s (sd = 7.42 s) and the mean delay in the SCI group being 8.20 s (sd = 8.80 s). Similarly, for HbO concentration in the right inferior parietal lobe, we observed slightly higher CVR delay values in the AB group compared to that of the SCI group (linear mixed-effects model, fixed-effects estimate = –6.136, Sattherthwaite’s t-test, t = – 2.332, p = 0.020) (Table 3). No significant differences in delays were observed between the SCI and AB groups for HbT concentration across all ROIs (Satterthwaite’s t-tests, p > 0.05). The CVR delays varied across the different ROI investigated; however, since we were primarily interested in differences in CVR delays between the AB and SCI groups, statistical results from the linear mixed-effects models were only reported for group differences in each region (Table 3).

**Figure 5.**
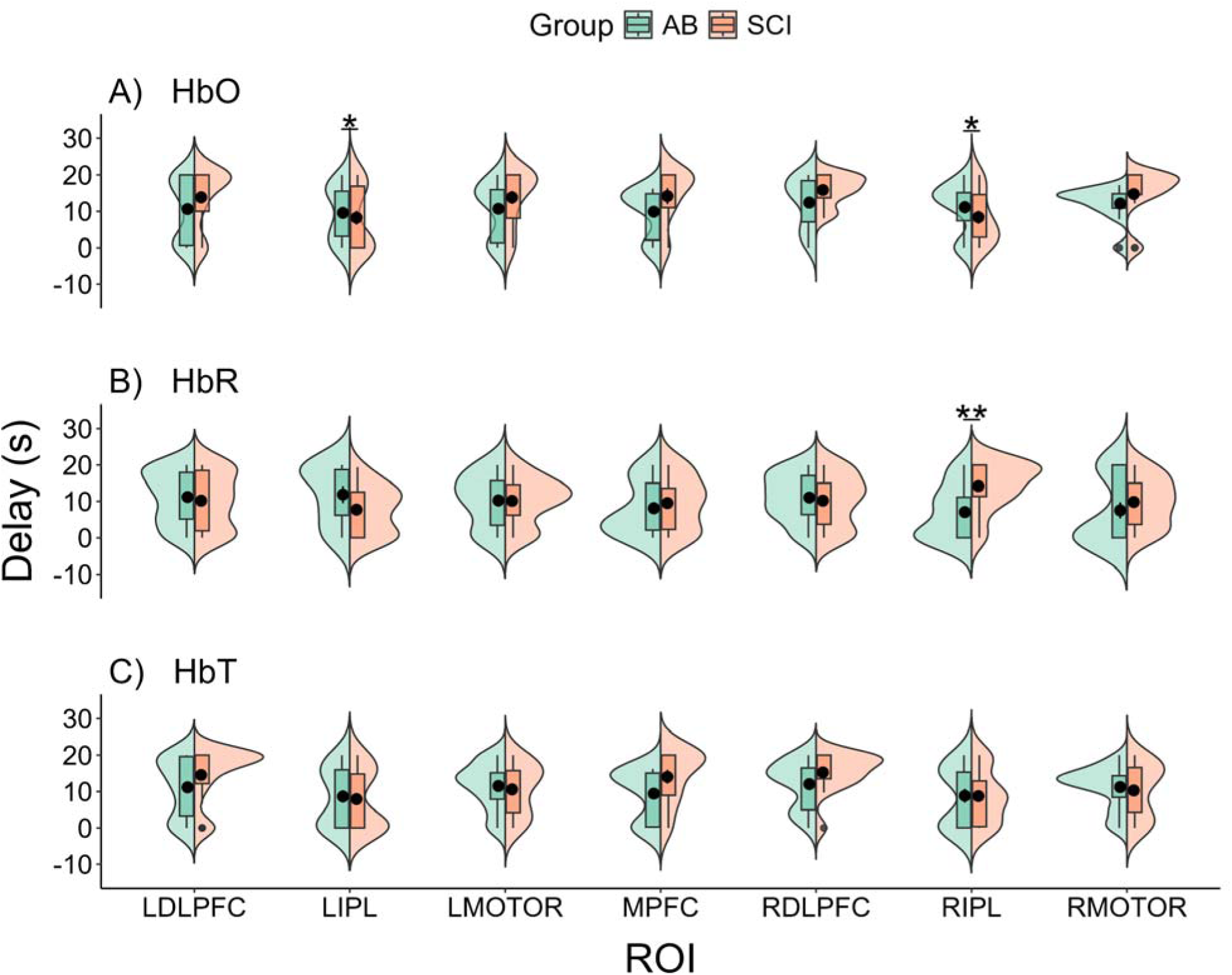
Breath-hold CVR delay values between AB and SCI groups for. **A)** oxy-hemoglobin (HbO), **B)** deoxy-hemoglobin (HbR), and **C)** total hemoglobin (HbT), across the left dorsolateral prefrontal cortex (LDLPFC), left inferior parietal lobe (LIPL), left motor cortex (LMOTOR), medial prefrontal cortex (MPFC), right dorsolateral prefrontal cortex (RDLPFC), right inferior parietal lobe (RIPL), and right motor cortex (RMOTOR). The black circles indicate the mean of the CVR delays and the distributions of the delays are shown for the AB and SCI groups, denoted by green and orange, respectively.

**Table 3.** Mixed effects model results for CVR delay comparisons between SCI and AB groups, across each ROI. One ROI is left out in the reports (the LDLPFC region) due to being the reference level for all comparisons.

| ROI | Estimate |  |  | Standard Error |  |  | t-value |  |  | p-value |  |  |
| --- | --- | --- | --- | --- | --- | --- | --- | --- | --- | --- | --- | --- |
|  | HbO | HbR | HbT | HbO | HbR | HbT | HbO | HbR | HbT | HbO | HbR | HbT |
| LIPL | -6.027 | -2.547 | -3.919 | 2.394 | 2.768 | 2.254 | -2.518 | -0.920 | -1.739 | 0.012 * | 0.358 | 0.083 |
| LMOTOR | -0.218 | -1.318 | -3.885 | 2.390 | 2.509 | 2.299 | -0.091 | -0.525 | -1.690 | 0.927 | 0.600 | 0.092 |
| MPFC | 2.313 | 1.838 | 3.723 | 2.539 | 2.433 | 2.449 | 0.911 | 0.756 | 1.520 | 0.363 | 0.450 | 0.129 |
| RDLPFC | 0.653 | -0.276 | 0.483 | 2.145 | 1.930 | 2.107 | 0.304 | -0.143 | 0.229 | 0.761 | 0.887 | 0.819 |
| RIPL | -6.136 | 6.565 | -3.254 | 2.631 | 2.465 | 2.338 | -2.332 | 2.663 | -1.392 | 0.020 * | 0.008 ** | 0.165 |
| RMOTOR | -0.425 | 2.681 | -2.803 | 2.444 | 2.504 | 2.303 | -0.174 | 1.071 | -1.217 | 0.862 | 0.285 | 0.224 |

### 3.3. CVR Association with Duration Since Injury and Injury Level

Upon further investigation of the right inferior parietal region’s CVR delays, we found a significant association between the CVR delay and duration since injury for HbT concentration values in the right inferior parietal lobe. As the duration since injury increased, the CVR delay values decreased (Pearson’s r correlation, r = –0.59, p = 0.04) (Figure 6A). For HbO and HbR concentration values, no significant associations were found between the mean CVR delays in the right inferior parietal lobe and the duration since injury (Pearson’s r correlation, HbO: r = – 0.09, p = 0.82, HbR: r = 0.28, p = 0.41). Similarly, no significant associations were found between the CVR delays in the right inferior parietal lobe and the injury level score for either HbO, HbR, or HbT (Pearson’s r correlation, p > 0.05) (Figure 6B). For subjects without sufficient data in any of the channels in the right inferior parietal lobe (channels 13, 14, 17, and 18), the data were not included in the correlational analyses.

**Figure 6.**
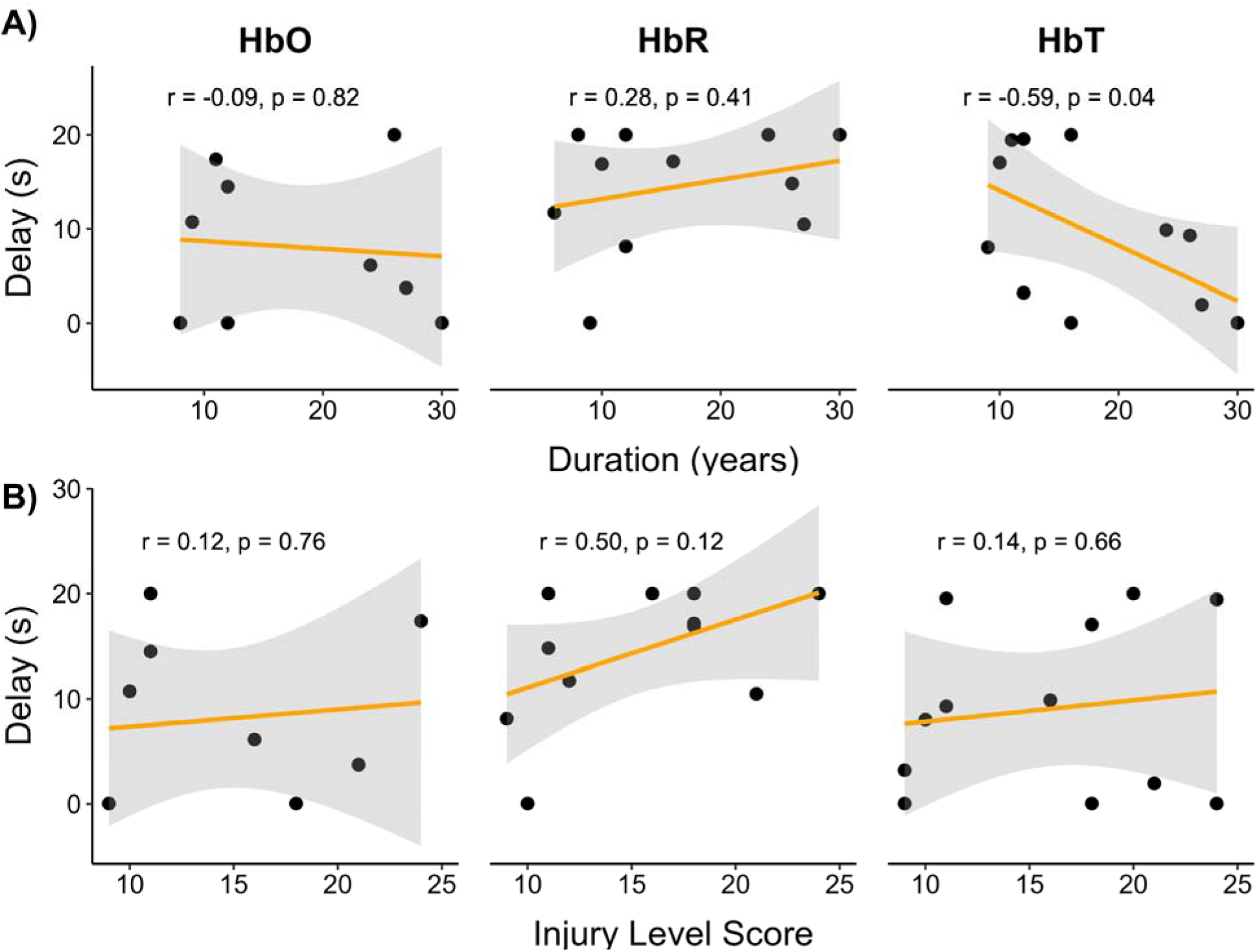
Association between the mean CVR delays in the right inferior parietal lobe and the. **A)** duration since injury (years) and **B)** spinal injury level score in the SCI group, for oxygenated hemoglobin (HbO), deoxygenated hemoglobin (HbR), and total hemoglobin (HbT) concentrations. Pearson’s correlation was performed and a least-squares line is shown in orange with the standard error of the mean in gray.

### 3.4. Variability in CVR Across the Brain

The SCI group showed significantly higher variability of CVR delays for HbO concentration compared to the AB group, as calculated by the standard deviation of CVR values across the brain (across all channels) (two-sample independent t-test, p = 0.0168) (Figure 7B). For CVR amplitude, there were no significant differences in variability across the brain between the AB and SCI groups across HbO, HbR, and HbT concentrations (independent-samples t-test, p > 0.05) (Figure 7A). For CVR amplitude, the whole-brain variability ranged from 0.15 to 0.25 for both AB and SCI groups, showing minimal differences in variability.

**Figure 7.**
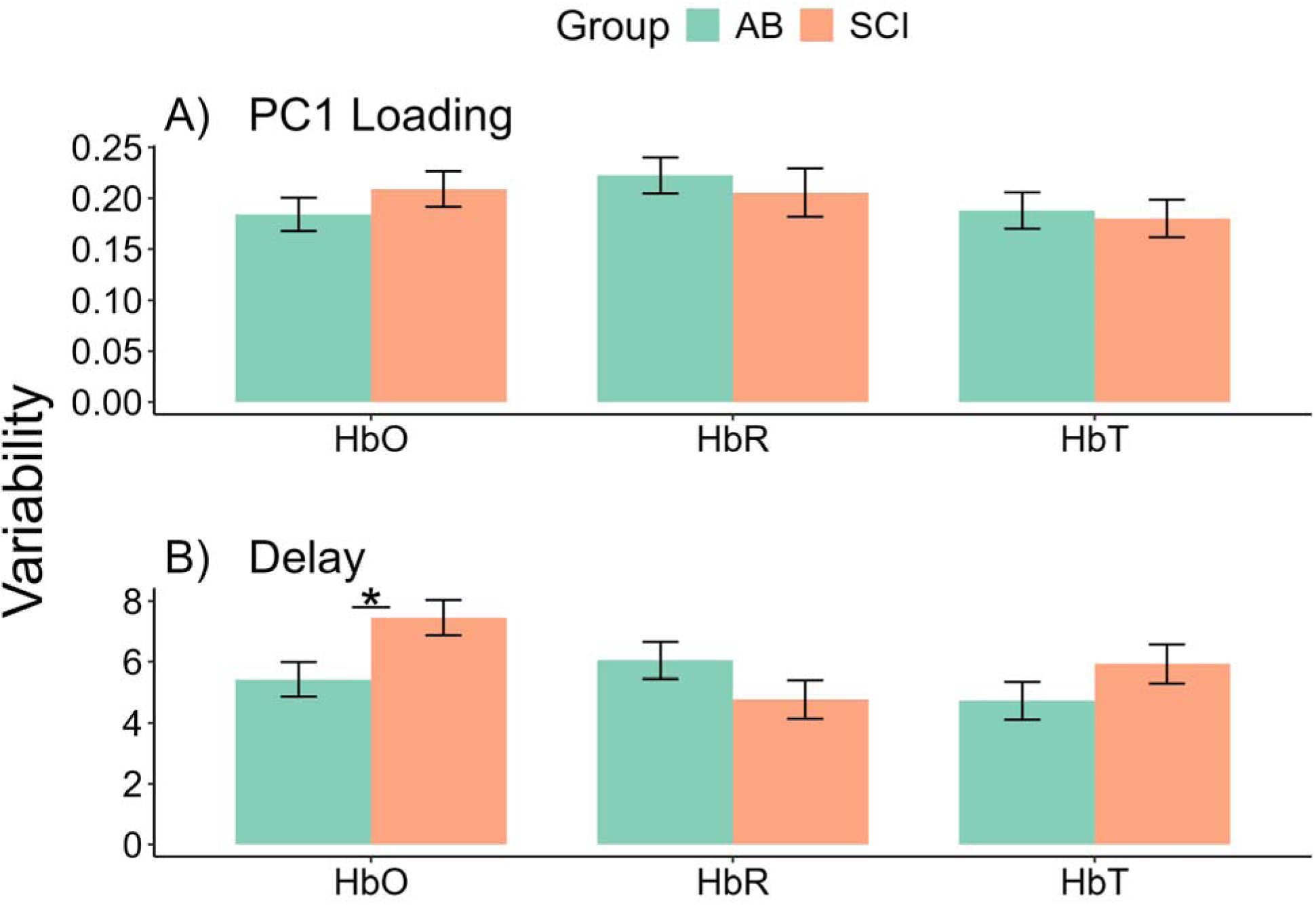
Variability in breath-hold CVR. **A)** amplitude (PC1 Loading) and **B)** delays across the brain between the AB and SCI groups, shown in green and orange, respectively. For HbO data, we found significant differences between the AB and SCI groups in the variability of delays across the brain (across channels) (two-sample t-test, p = 0.0168). Error bars indicate ±1 standard error of the mean.

## 4. Discussion

In this study, we examined differences in the CVR amplitude and delays between SCI and AB individuals using fNIRS during a hypercapnic breath-holding task. We also investigated the association between CVR components and injury characteristics in individuals with SCI. We report significantly delayed CVR in the right inferior parietal lobe in individuals with SCI compared to AB individuals, while the amplitude of CVR was unchanged. CVR also showed different characteristics between HbO, HbR, and HbT concentrations. As the duration since injury increased for individuals with SCI, CVR delay in the right inferior parietal lobe was found to decrease. To the best of our knowledge, this is the first study to report differences in CVR delays between SCI and AB individuals quantified by breath-hold fNIRS and cross-correlation analysis.

### 4.1. No differences in CVR amplitude between the SCI and AB groups

When comparing CVR amplitude between SCI and AB groups, we observed no significant differences between the two groups. This is consistent with Weber and colleagues’ results that used fMRI to quantify CVR, in which no significant differences were observed in the standalone and steady-state CVR results between SCI and AB groups (Weber et al., 2022). They argue that unchanged full CVR may be masked by increased CVR in individuals with thoracic injuries and decreased CVR in those with cervical injuries. Furthermore, Wilson and colleagues reported that CVR is maintained in individuals with tetraplegia, whereas dynamic cerebral autoregulation may be altered, suggesting adaptive mechanisms in those with chronic tetraplegia (Wilson et al., 2010). Although our CVR metric was quantified using a hypercapnic breath-hold stimulus, the previous studies investigating CVR in individuals with SCI used CO2 gas inhalation and may be limited by the small sample sizes (7 and 6 SCI participants, respectively) (Weber et al., 2022; Wilson et al., 2010). Despite these differences, our results on the time-independent CVR metric are consistent with their conclusions. An advantage of our method to quantify CVR amplitude by using PCA is that there is no biased assumption on the specific HRF shape associated with the breath-hold CVR response, thus providing a time-independent and HRF-shape independent metric of CVR.

### 4.2. Individuals with SCI have significantly longer CVR delays than AB individuals

We found significantly longer CVR delays in the SCI group compared to the AB group in the right inferior parietal lobe. Longer delays in CVR is associated with poorer vascular health (K. B. Miller et al., 2019; Sobczyk et al., 2021; Stringer et al., 2021), suggesting reduced vascular health in the right inferior parietal lobe in individuals with SCI. The right inferior parietal lobe is involved in visuospatial attention and sensorimotor integration, in addition to executive function (Corbetta & Shulman, 2002; Numssen et al., 2021). Delays in this region may be due to loss of sensorimotor function in individuals with SCI after injury, particularly for the paraplegic cohort recruited in the present study. However, it is important to note that we did not find significant differences in the CVR amplitude despite finding differences in the CVR delays. This may suggest a compensatory mechanism in which a longer CVR delay in individuals with SCI is required for comparable CVR amplitudes to that of the AB group. Furthermore, a significant delay in CVR in the SCI group was observed for HbR only, which may reflect more sensitive changes to a breath-hold task in HbR rather than to HbO concentrations in the blood. CVR quantified using blood-oxygen-level-dependent (BOLD) fMRI techniques cannot fully capture differences in CVR dynamics between the different hemoglobin species. This is due to HbR being paramagnetic, causing dephasing and shortening of the transverse relaxation time (T_2_*) weight, which decreases the fMRI BOLD signal; therefore, a decrease in HbR subsequently increases the strength of the signal (Mcintyre et al., 2003). HbO on the other hand, is not paramagnetic and has very little contribution to relaxation rates. Therefore, using fNIRS, we can better distinguish between CVR measured by dynamic changes not only in HbR, but also in that of HbO and HbT. This shows a unique characteristic of quantifying CVR using fNIRS as compared to that of fMRI, which relies primarily on HbR changes. Future studies may want to further investigate the different sensitivities of HbO, HbR, and HbT in quantifying CVR using fNIRS.

### 4.3. CVR delays in the right inferior parietal lobe is associated with the duration since injury

We further investigated the CVR delays in the right inferior parietal lobe in individuals with SCI and found a significant association between CVR delays with the duration since injury, for HbT concentration. As the duration since injury increased for the SCI group, the CVR delays decreased. This suggests that over time, the cerebrovascular health of individuals with SCI may improve, as the cerebrovasculature remodels and motor region connections reorganize after injury (Kokotilo et al., 2009). These improvements in CVR delays in the right inferior parietal lobe may also be related to the frequency of rehabilitation or exercise programs that individuals may be enrolled in. Although it is not clear the extent to which individuals with SCI in this cohort actively participate in physical rehabilitation training, generally, exercise improves cerebrovascular health, as blood perfusion is increased in the brain (Bliss et al., 2021; Davenport et al., 2012).

### 4.4. Variability in CVR across the brain

We found significantly higher variability in the delay of CVR across the brain in individuals with SCI compared to AB individuals, for HbO concentration. This may be explained by the differences in injury characteristics in the SCI group, whereas those in the AB group were more likely to be more homogeneous. As injury characteristics such as the completeness of injury, level of injury, duration since injury, and cause of injury differ across individuals in the SCI group, it is consistent that the CVR characteristics across the brain would also differ more in the SCI group. In healthy aging subjects, CVR has been shown to differ across brain regions, with CVR in particular regions such as the hippocampus being associated with memory scores (Catchlove et al., 2018). The heterogeneous nature of CVR has been reported in previous studies (D. Y. Chen, Di, & Biswal, 2024; D. Y. Chen, Di, Yu, et al., 2024; Kastrup et al., 1999; Moia et al., 2020; van Niftrik et al., 2023); however, it is unclear whether a measure of the heterogeneity of CVR may reveal vascular pathologies in the brain, particularly in patient populations.

### 4.5. Limitations

The present study contains a lack of physiological measures such as end-tidal CO2 measures and respiratory belt data. However, it is important to note that the use of a respiratory belt on patient populations such as those with SCI may be too constricting and may not be comfortable to wear while performing a breath-holding task. It is important that participants were as comfortable as possible when performing the breath-holding tasks, thus although extra physiological sensors would be helpful, it may cause other unwanted side effects from the patients. Future studies may also want to look at the differences in CVR in individuals with thoracic vs. cervical injuries and those who have tetraplegia vs. paraplegia, since autonomic dysfunction can vary greatly based on the severity of injury. Furthermore, future studies may also want to look into using breath-hold CVR as a regressor in fNIRS studies to account for possible differences between patient populations and controls that may be due primarily to vascular differences rather than neuronal differences.

## 5. Conclusion

Using fNIRS and a breath-holding hypercapnic task, the present study investigated differences in CVR between individuals with SCI and AB controls. We found significantly delayed CVR in the right inferior parietal lobe in individuals with SCI compared to AB controls, and no significant differences in the amplitude of CVR between the SCI and AB groups. The delays in CVR in the SCI group were also associated with the duration since injury, in which a longer duration since injury resulted in a shortened delay in CVR. This suggests adaptive CVR mechanisms with time as individuals with SCI recover and undergo functional brain reorganization.

## CRediT authorship contribution statement

**Donna Y. Chen:** conceptualization, methodology, software, formal analysis, investigation, data curation, writing – original draft. **Xin Di:** conceptualization, supervision, writing – review&editing. **Keerthana Deepti Karunakaran:** writing – review&editing. **Hai Sun:** writing – review&editing. **Saikat Pal:** resources, writing – review&editing. **Bharat Biswal:** conceptualization, supervision, funding acquisition, writing – review&editing.

## Data Availability

All data produced in the present study are available upon reasonable request to the authors

## Acknowledgements

The authors would like to sincerely thank Dr. Christopher Cirnigliaro, Ms. Annie Kutlik, and Mr. Steven Knezevic from the Spinal Cord Damage Research Center, James J. Peters Veterans Affairs Medical Center, Bronx, NY for helping with patient recruitment for this study. Research reported in this publication was supported by the New Jersey Commission on Spinal Cord Research under Award Number CSCR22ERG026 to B.B.B. This work was also supported by the National Center for Advancing Translational Sciences (NCATS) of the National Institutes of Health (NIH) under Award Number TL1TR003019 to D.Y.C. The content is solely the responsibility of the authors and does not necessarily represent the official views of the funding agencies.

